# Cortico-striatal engagement during cue-reactivity, reappraisal, and savoring of drug and non-drug stimuli predicts craving in heroin addiction

**DOI:** 10.1101/2022.05.27.22275628

**Authors:** Yuefeng Huang, Ahmet O. Ceceli, Greg Kronberg, Sarah King, Pias Malaker, Nelly Alia-Klein, Eric Garland, Rita Z. Goldstein

## Abstract

**Importance:** Heroin addiction is rampant and persistent, with devastating consequences to the public health, necessitating further study into the neurobiological mechanisms of drug cue-reactivity and craving-reducing interventions (e.g., reappraisal and savoring).

**Objective:** To document cortico-striatal reactivity during passive viewing, reappraisal, and savoring, as predictors of heroin craving in individuals with heroin use disorder (iHUD) vs. controls.

**Design:** A cross-sectional study (11/2020-09/2021), with a novel fMRI task in iHUD vs. controls.

**Setting:** iHUD and controls were recruited from treatment facilities and surrounding neighborhoods, respectively.

**Participants:** iHUD (N=32) [40.3±8.8 years; 7 (21.9%) women] and age-/sex-matched controls (N=21) [40.6±10.8 years; 8 (38%) women].

**Main Outcomes and Measures:** Between-group blood-oxygen-level-dependent signal differences during cue-reactivity/reappraisal/savoring and their direct contrast (reappraisal vs. savoring), and correlations with drug craving in iHUD.

**Results:** Drug cue-reactivity (look drug>neutral) revealed higher nucleus accumbens and ventromedial prefrontal cortical activity in iHUD vs. controls (Z>3.1, p<.05), the latter positively correlated with post-task drug cravings (r^2^=.47, p<.001). In contrast, controls showed higher dorsolateral prefrontal cortex (dlPFC) and inferior frontal gyrus (IFG) reactivity to food cues (>drug; Z>3.1, p<.05; with the opposite pattern for the iHUD). Both drug-reappraisal and food-savoring (vs. respective passive viewing) showed increased activity in the IFG and supplementary motor area in all participants; the higher the dlPFC/IFG drug reappraisal in iHUD, the lower the post-MRI drug cue-induced craving (Z>3.1, p<.05). A direct comparison (drug-reappraisal vs. food-savoring) revealed higher drug-reappraisal in the ventral caudate and PFC regions in the iHUD (Z>3.1, p<.05), as predicted in the striatum by pre-task drug cravings (Z>2.57, p<.05); in controls, these areas showed instead higher food-savoring (Z>3.1, p<.05).

**Conclusions and Relevance:** We demonstrate upregulated cortico-striatal activity during drug-cue exposure (while passively looking or reappraising) and impaired reactivity during processing (looking or savoring) of non-drug rewards in heroin addiction. These results bolster the impaired response inhibition and salience attribution model of drug addiction, previously supported mostly by results in stimulant addiction. Normalizing cortico-striatal function by reducing drug cue-reactivity (e.g., reappraising drug cues) and enhancing natural reward valuation (e.g., savoring food stimuli) may inform therapeutic mechanisms for reducing drug craving/seeking in heroin addiction.

**Key Points:** *Question:* What are the cortico-striatal brain regions driving reactivity to and reappraisal of drug vs. savoring food cues in individuals with heroin use disorder (iHUD); do these activations contribute to self-reported drug cravings?

*Findings:* In this cross-sectional study, and compared to matched healthy control subjects, iHUD exhibited cortico-striatal reward and self-control regional hyperactivations to drug cues during both passive viewing of pictures and their reappraisal as compared to savoring of alternative reward (images of food); and these activations correlated with drug cravings.

*Meaning:* We identified drug-cue biased processing in cortico-striatal regions in iHUD, providing potential therapeutic biomarkers for neuromodulation and cognitive training in reducing hyper-responsiveness to drug cues and enhancing responses to alternative natural rewards.

## Introduction

Drug overdoses claimed 100,000+ lives in 2021, 75,000+ of which were opioid-related.^1^ Despite the tremendous cost of opioid addiction, little is known about its underlying neurobiological mechanisms.^2^ The impaired response inhibition and salience attribution (iRISA) model posits that addiction is driven by enhanced salience of drug cues at the expense of non-drug reinforcers with a concomitant decrease in self-control.^3,4^ Literature reviews of iRISA neuroimaging studies in drug addiction reveal hyperactivations during drug cue exposure in cortico-striatal regions underlying reward processing [e.g., nucleus accumbens (NAcc), ventromedial PFC (vmPFC), and orbitofrontal cortex (OFC)],^5–7^ salience attribution [e.g., NAcc, dorsal anterior cingulate cortex (dACC), and insula],^8–10^ and executive function [e.g., dorsolateral PFC (dlPFC) and inferior frontal gyrus (IFG)]^6,11,12^—as associated with craving.^13,14^ During non-drug cues processing (including when performing inhibitory control, decision-making, and social-emotional tasks), these regions/networks exhibit hypoactivations.^13^ However, few functional neuroimaging studies have included individuals with heroin use disorder (iHUD) (e.g., 6 of the 107 studies reviewed in Zilverstand et al.,^13^), precluding generalization of the iRISA predictions and identification of treatment mechanisms to this population.

The underlying neural mechanisms of drug cue-reactivity^15^ are typically mapped while participants passively view drug and non-drug stimuli during functional MRI (fMRI). These mechanisms involve the recruitment of the NAcc and vmPFC/OFC in response to drug vs. neutral cues in individuals with alcohol,^16,17^ nicotine,^18,19^ cannabis,^20^ or cocaine^21–23^ use disorders. Increased drug cue related activity in these cortico-striatal regions is associated with higher self-reported craving in alcohol,^24^ nicotine,^18^ and cannabis^20^ addiction. Similarly in iHUD and compared to healthy control subjects (HC), passively viewing drug vs. neutral cues elicited enhanced activity in the NAcc and vmPFC/OFC among other nodes of the dopaminergic reward network,^25–29^ as associated with heroin craving^25,26^ and predictive of subsequent relapse.^25^ Additionally, within iHUD and compared to viewing a non-drug salient reinforcer (sexual cues), viewing drug cues increased dlPFC and IFG activity.^30^ However, most of these cue-reactivity studies in iHUD lacked a HC group and primarily focused on a drug vs. neutral comparison (and not to another salient reinforcer), limiting a thorough inspection of the relevance of the iRISA model to heroin addiction.

Reducing drug cue-reactivity has been a target of numerous interventions in addiction.^31^ In the general population, both emotional downregulation and its upregulation commonly activate the dlPFC and IFG, with both emotional regulation approaches also showing distinct activation patterns (e.g., the former decreases activity in regions receiving interoceptive input and the latter increases activity in regions associated with emotional experience, such as the striatum and ACC).^32^ Similarly, two common interventions to reduce craving encompass the deployment of PFC-mediated cognitive control (akin to emotional downregulation) and the deployment of subcortically mediated savoring (akin to emotional upregulation).^33,34^ An example of the former is cognitive reappraisal, a regulatory strategy that involves re-interpreting the meaning of an emotionally salient cue, driven by PFC-mediated attentional reorienting^35^, goal planning and maintenance,^11^ and cognitive flexibility.^36^ Meta-analyses report increased dlPFC, IFG, dACC, and supplementary motor area (SMA) activations during reappraisal of negative stimuli in the general^37–39^ and clinical populations.^40^ Recent research identified opioid misuse-related deficits in reappraisal of negative emotional stimuli as indicated by the late positive potential (LPP).^41^ In contrast to some evidence in individuals with cocaine addiction^42^. There is only indirect evidence for the effects of cognitive reappraisal on drug cue-reactivity in opiate addiction and misuse. For example, behavioral therapies such as Mindfulness-Oriented Recovery Enhancement (MORE)^43^ include reappraisal training as part of their regimen. These therapies are associated with neurophysiological down-regulation of emotional reactivity in opioid misusing chronic pain patients, as assayed via the LPP during reappraisal vs. looking at opioid cues,^44^ and as related to reduced opioid use, craving, and emotional distress at a nine-month follow-up.^45^

An example of the latter, another potential therapeutic mechanism suggested by the iRISA model, involves savoring, or increasing the salience of non-drug alternative reinforcers (e.g., food and pleasant images). In individuals with cocaine addiction, longer abstinence duration was associated with recovery of such hedonic processing as assessed with LPP responses to non-drug pleasant images and correlated with decreased craving.^46^ Clinically, savoring is incorporated in MORE,^43^ where participants are trained to attend to and relish natural rewards, an active strategy to increase the salience of non-drug cues. In the general population, savoring positive autobiographical memories increases cortico-striatal blood-oxygen-level-dependent (BOLD) activity, particularly in the striatum and the vmPFC.^47^ Compared to HC, smokers who underwent savoring training as part of MORE increased NAcc and vmPFC activity while savoring positive emotional images, as associated with reduced cigarette smoking.^48^ In chronic pain patients treated with MORE, savoring was associated with increased centroparietal LPP during exposure to natural reward cues, increased positive affect, and decreased prospective anhedonia and opioid craving.^44,49,50^

To the best of our knowledge, no study to date has investigated the cortico-striatal mechanisms underlying cue-reactivity, reappraisal, and savoring, and their putative association with drug craving, in iHUD as compared to HC. To this end, our novel fMRI paradigm captured the brain bases of reactivity to and reappraisal of drug cues and savoring of non-drug reward cues (images of food), exploring their associations with ratings of drug craving, in medically-stabilized inpatients with HUD. Because both reappraisal and savoring engage common (and distinct) neural pathways, potentially competing for the same resources, here for the first time we directly compared both drug cue-reactivity reduction strategies in iHUD. Following predictions from iRISA, comparing iHUD to HC, we hypothesized 1) increased engagement of the cortico-striatal regions across the reward, salience, and control networks in response to drug cues for passive viewing (vs. neutral or food cues) but also during reappraisal as directly compared to savoring; 2) decreased cognitive control network activations while reappraising (vs. looking at) drug cues; 3) decreased reward and cognitive control network activations while savoring (vs. looking at) food cues; and 4) correlations between these activation patterns with cravings in the iHUD.

## Methods

### Participants

Thirty-two treatment-seeking iHUD (mean age = 40.3±8.8, 7 women) from a medication-assisted inpatient treatment facility and 21 age-and sex-matched HC (mean age = 40.6±10.8, 8 women) from the surrounding community (via flyers and local advertisements) were recruited for the study (see Table 1 for details). All participants reviewed the study procedures and provided written informed consent. The study was approved by the Icahn School of Medicine at Mount Sinai’s Institutional Review Board.

**Table 1.** Sample profile.

|  | HUD (N = 32) | HC (N = 21) | Significance test |
| --- | --- | --- | --- |
| Age <sup>a</sup> | 40.25 ± 8.82<br>Min: 27.7, Max: 60.3 | 40.58 ± 10.84<br>Min: 25.9, Max: 58.5 | $W=334, p=.978$ |
| Sex (M/F) <sup>b</sup> | 25/7 | 13/8 | $\chi^2(1, N=53)=0.94, p=.332$ |
| Race (Black/White/Other & mixed/Missing) <sup>b</sup> | 2/22/4/4 | 8/11/2/0 | $\chi^2(2, N=53)=7.08, p=.030$ |
| Ethnicity (Hispanic/Non-Hispanic) <sup>b</sup> | 14/18 | 3/18 | $\chi^2(1, N=53)=3.78, p=.052$ |
| Handedness (Left/Right) <sup>b</sup> | 5/27 | 4/17 | $\chi^2(1, N=53)<.001, p=1$ |
| Education (years; 12=high school grad) <sup>a</sup> | 12.25 ± 2.29<br>Min: 9, Max: 17 | 16.76 ± 2.84<br>Min: 13, Max: 25 | $W=604.5, p<.001^*$ |
| Verbal IQ [Wide Range Intelligence Test (WRAT)-III standard score] <sup>c</sup> | 94.03 ± 12.17<br>Min: 71, Max: 116 | 110.10 ± 7.71<br>Min: 93, Max: 123 | $t(50.97)=5.90, p<.001^*$ |
| Non-verbal IQ [Wechsler Abbreviated Scale of Intelligence (WASI) matrix reasoning scaled score] <sup>a</sup> | 10.74 ± 2.71<br>Min: 3, Max: 15,<br>Missing: 1 | 11.71 ± 3.33<br>Min: 4, Max: 17 | $W=405.5, p=.135$ |
| Beck Depression Inventory-II <sup>a</sup> | 13.7 ± 12.3<br>Min: 0, Max: 46,<br>Missing: 1 | 3.38 ± 4.67<br>Min: 0, Max: 17 | $W=121.5, p<.001^*$ |
| Smoker (never/past/current) <sup>b</sup> | 0/0/32 | 17/3/1 | $\chi^2(2, N=53)=48.95, p<.001^*$ |
| Age of onset (first use) | 25.06 ± 7.48<br>Min: 15, Max: 42 | - | - |
| Age of onset (regular use) | 25.88 ± 7.37<br>Min: 15, Max: 43 | - | - |
| Period of heaviest use (years) | 4.69 ± 6.04<br>Min: 0.08, Max: 27,<br>Missing: 1 | - | - |
| Heroin lifetime use (years) | 10.98 ± 7.66<br>Min: 2, Max: 27 | - | - |
| Last time use heroin (days) | 196.78 ± 255.04<br>Min: 7, Max: 1007 | - | - |
| Heroin Severity of Dependence Scale (SDS) | 10.44 ± 3.64<br>Min: 0, Max: 15 | - | - |
| Heroin Craving Questionnaire (HCQ) | 42.84 ± 15.85<br>Min: 16, Max: 86,<br>Missing: 1 | - | - |
| Short Opiate Withdrawal Scale (SOWS) | 3.16 ± 3.37<br>Min: 0, Max: 12 | - | - |

Significant group differences after controlling for multiple comparisons ( $p<.05/10$ ) are flagged with an asterisk. Continuous variables are reported as mean ± standard deviation. <sup>a</sup> Wilcoxon rank-sum test; <sup>b</sup> Chi-squared test; <sup>c</sup> two-sample t-test with unequal variance. In total 7 iHUD have missing data for race/WASI/BDI/HCQ/period of heaviest use.

A comprehensive clinical diagnostic interview was conducted by trained research staff under a clinical psychologist’s supervision, encompassing the Mini-International Neuropsychiatric Interview^51^ and Addiction Severity Index^52^, a semi-structured instrument capturing drug use history and severity. Severity of drug dependence, craving, and withdrawal symptoms were assessed using the Severity of Dependence Scale,^53^ the Heroin Craving Questionnaire (a modified version of the Cocaine Craving Questionnaire)^54^ and the Subjective Opiate Withdrawal Scale,^55^ respectively. A brief physical examination including heart rate, blood pressure, urine drug toxicology, breath alcohol, and carbon monoxide levels, and a review of medical history were also performed by trained research staff.

All iHUD met DSM-5 criteria for current heroin use disorder and were stabilized on medication (methadone and/or buprenorphine) as confirmed by urine assays (methadone=26, buprenorphine=5, methadone & buprenorphine=1); four participants used heroin 30 days within the study (average days of heroin use in the past month = 0.22±0.75). Urine toxicology results were negative for all 21 HC participants. Primary routes of heroin administration within iHUD included 17 intravenous, 11 intranasal, 3 smoking/inhaling, and 1 oral. All 32 iHUD, and one HC participant, were current cigarette smokers [Fagerström Test for Nicotine Dependence (FTND) total score in the iHUD=3.69±1.53, number of cigarettes smoked per day=2.33±2.20] (see supplement for exclusion criteria and comorbidities).

### Experimental Design and Statistical Analysis

#### fMRI task paradigm

Participants viewed drug, food, and neutral images in a novel block-design fMRI cue-reactivity task. The images were collected from freely available online sources (e.g., social media, Google image search). Drug images included diverse individuals using heroin via injection, sniffing, smoking, or engaging with heroin-related paraphernalia. Images including faces were cropped below the nose to avoid potential effects of reactivity to facial expressions. Food and neutral images were visually matched to the drug images on body/non-body part ratio and brightness. These images were presented under three separate instruction conditions in randomized order (Figure 1). In the “look” condition, participants were instructed to passively look at the three types of images. In the “reappraise” condition, they were instructed to actively down-regulate their emotional reactivity to the drug images. In the “savor” condition, they were instructed to actively up-regulate their emotional reactivity to food images (see supplement for verbatim task instructions). Participants were informed of the task conditions via a horizontal flat line, down arrow, or up arrow above the images for the look, reappraise, and savor conditions, respectively. Sixteen-second “on” blocks with four images of the same cue type were displayed for 4 seconds each, followed by a 10-second “off” block for a total of 9 blocks for the “look” (3 blocks for each of the three cue types), and 3 blocks for each of the “savor” and “reappraise” conditions per each of the three task runs, totaling approximately 20 minutes. Participants practiced these instructions outside of the scanner in an abbreviated version of the task. Immediately before and after the task, participants provided self-reported 10-point (0-9) drug and food craving and task motivation ratings, and self-evaluation of difficulty and success of reappraisal and savoring after the task. After the MRI session, each subject provided 5-point (1-5) valence, arousal, and craving (cue-induced craving; for food and drug cues only) ratings on half of the drug, food and neutral images viewed during the task (see supplement for analyses).

**Figure 1:**
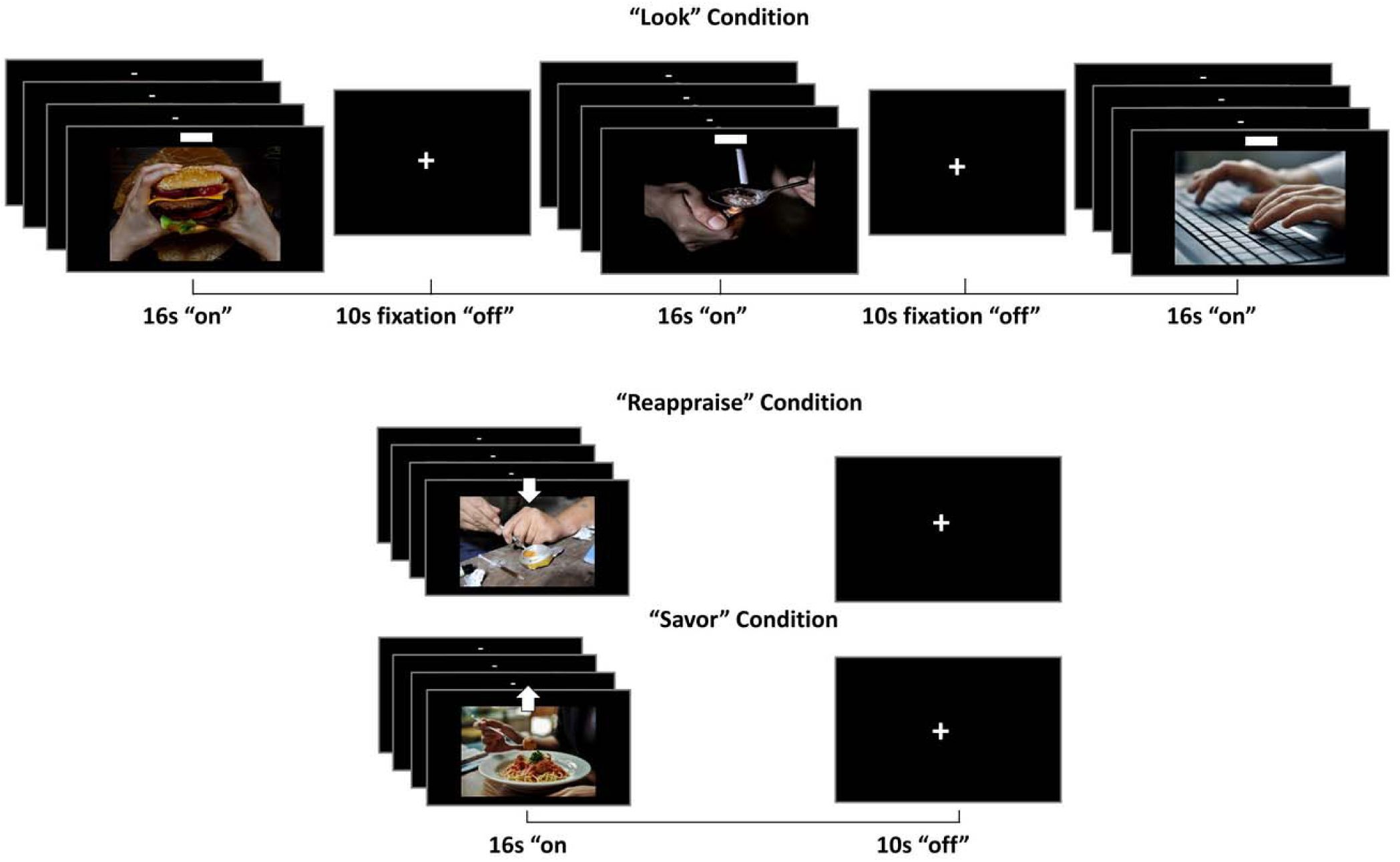
The fMRI cue-reactivity task. Participants were instructed to passively view 16-sec blocks of drug, food, and neutral images during the *look* condition, reduce emotional reactivity to drug, and increase emotional reactivity to food images during the *reappraise* and *savor* conditions, respectively. 10-sec off blocks separated each image block.

#### MRI data acquisition and preprocessing

Scanning was conducted on a Siemens 3T Skyra (Siemens Healthcare, Erlangen, Germany), using a 32-channel head coil (see supplement for scanning parameters). Raw BOLD-fMRI data were first converted to NIFTI using dcm2niix.^56^ Data were then preprocessed using the fMRIprep pipeline (version 20.2.1)^57^ (see supplement for summary).

#### BOLD-fMRI data analysis

The GLM regressors included three look events (drug, food, neutral), one reappraisal event (drug), and one savor event (food), sampled from the onset of the corresponding trials (16-sec length, convolved with a double gamma hemodynamic response function). The GLM also included the motion outlier time points as nuisance regressors. Fixation events were not modeled. The first-level analysis included the classic cue-reactivity contrasts (look drug/food>look neutral), and a drug cue-reactivity contrast controlling for a salient nondrug reward (look drug>look food). We also created contrasts representing reappraisal (reappraise drug>look drug and reappraise drug>savor food) and savoring (savor food>look food).

Parameter estimates from these contrasts were entered into a fixed-effects model to yield subject-level statistical maps, and group estimates were calculated in a higher-level analysis (separate t-tests comparing iHUD and HC on these contrasts) using FSL’s FLAME 1+2 (FMRIB’s Local Analysis of Mixed Effects) to improve group-level variance estimation and population inferences via Markov Chain Monte Carlo simulations.^58^ Whole-brain correlations were performed with general drug craving ratings (pre-, post-, and post-minus-pre) and drug cue-induced craving ratings using the post-MRI image ratings within iHUD. Unless specified otherwise, we used a cluster defining threshold of Z>3.1, corrected to a cluster-extent threshold of p<0.05, in line with common practices to minimize Type I error^59^ (see supplement for details).

## Results

### Participants

Groups differed significantly in education (HC>iHUD), verbal IQ (HC>iHUD), self-reported depression (iHUD>HC) and cigarette smoking (iHUD were all current smokers while HC were mostly never smokers) (p<.001; Table 1). These variables (including FTND scores/number of cigarettes smoked per day) were not significantly associated with any of our neuroimaging outcomes of interest and thus were not entered into fMRI analysis models as covariates.

### BOLD-fMRI Results

#### Cue-reactivity to drug and food images and correlation with drug cravings

Whole-brain between-group analysis of BOLD responses to drug cues (look drug>look neutral) revealed hyperactivations in the iHUD>HC in the left NAcc and right vmPFC as well as posterior cingulate cortex and other mostly parietal regions (Figure 2A & Table 2). Higher vmPFC (and OFC) drug cue-reactivity predicted post-task drug craving within iHUD (Figure 2B, Table 2). There were no significant group differences in response to food cues (look food>look neutral). When controlling for the potential contributions of salience (look drug> or <look food), iHUD exhibited drug-related hyperactivity in the right IFG and dlPFC among other mostly parietal and occipital regions, while the HC showed the opposite direction of results in the same PFC regions (Figure 2C, Table 2).

**Table 2.**
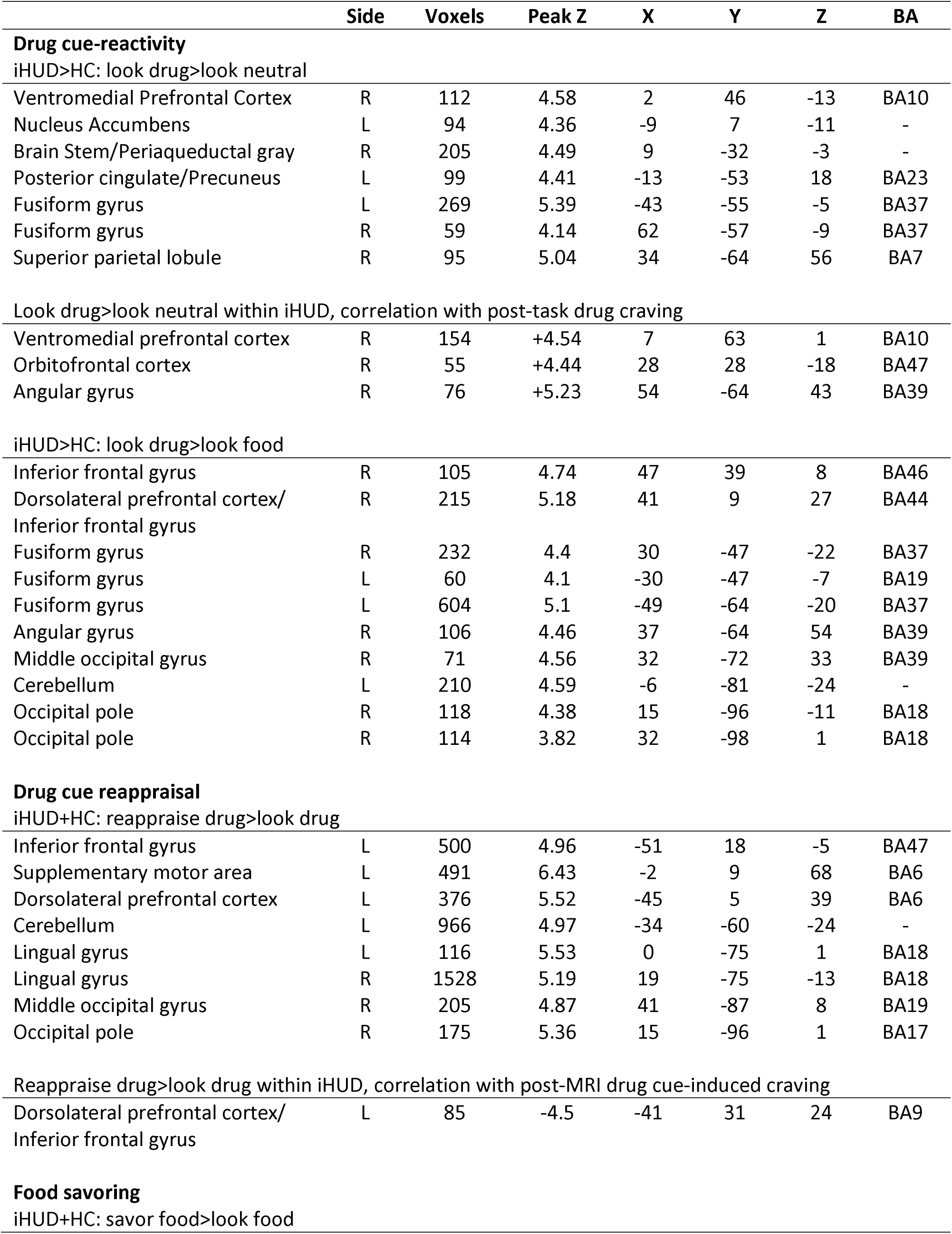
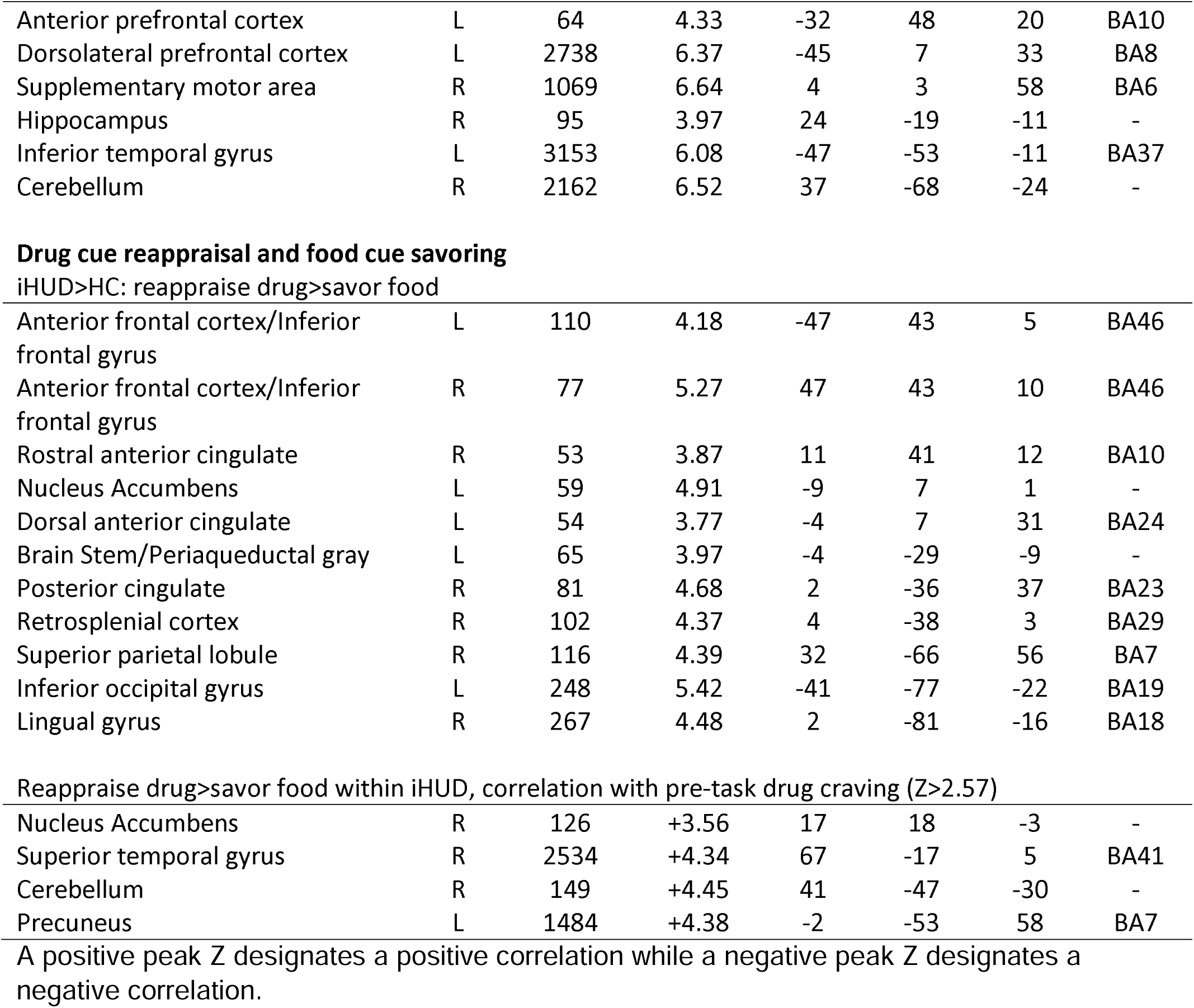
Coordinates for between-groups direct contrasts and within-group correlations.

**Figure 2:**
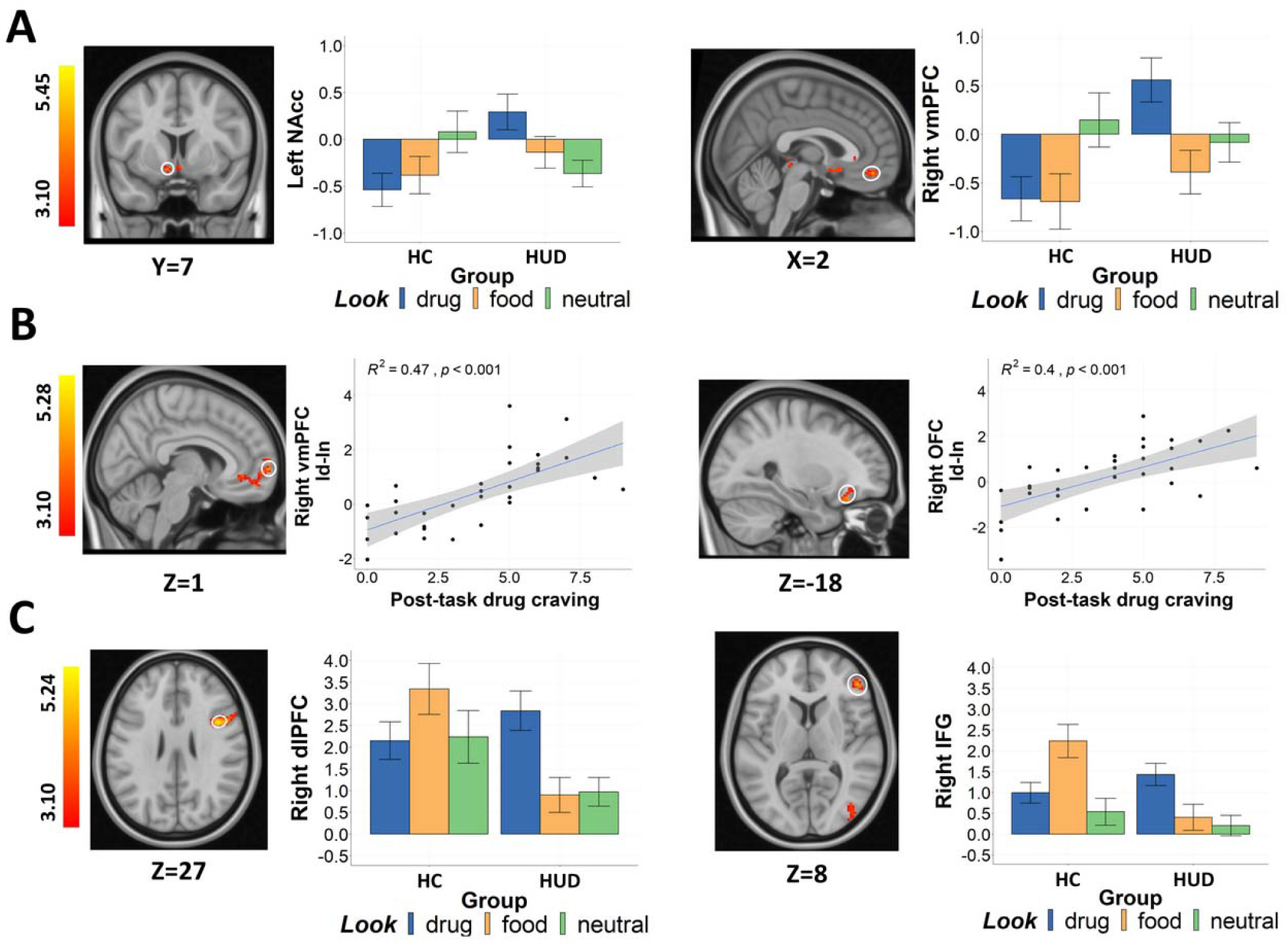
Drug cue-reactivity-related brain activity. A) Clusters indicating increased activity in iHUD compared to HC during the look drug>look neutral contrast in the left NAcc and right vmPFC. B) Clusters indicating significant whole-brain positive correlations between post-task drug craving and vmPFC (left panel) and OFC (right panel) activation during look drug>look neutral contrast within iHUD. C). Clusters indicating increased activity in the right dlPFC/IFG for iHUD compared to HC during the look drug>look food contrast. In all panels, for visualization purposes, bar graph and scatter plots depict BOLD signal and correlation derived via 3-mm radius masks centered on coordinates from peak activity (circled in white). Error bars represent standard error.

#### Drug cue reappraisal and food cue savoring and correlation with drug cravings

There were no significant group differences when comparing iHUD to HC during drug reappraisal (reappraise drug>look drug) and food savoring (savor food>look food); all participants had increased activity in the left IFG, left dlPFC, and bilateral SMA (Figure 3A & 3B, Table 2). During reappraisal (>look drug), higher left dlPFC/IFG activity predicted lower drug cue-induced craving within iHUD (Figure 3C Table 2). When controlling for salience/engagement and cognitive effort (reappraise drug>savor food), compared to the HC the iHUD exhibited increased activity in the bilateral IFG, left ventral caudate, left dACC and other regions such as the rostral ACC and posterior cingulate cortex (Figure 3D, Table 2). Using a lower threshold, higher pre-task baseline drug craving was associated with higher right NAcc activity during drug reappraisal>food savoring within iHUD (Z>2.57, *p*<0.05; Figure 3E, Table 2). See supplement (eFigure 3 eTable 1&2) for further inspection of these results accounting for passive viewing (reappraise drug minus look drug>savor food minus look food), and other within-iHUD correlations outside of our a priori regions of interest.

**Figure 3:**
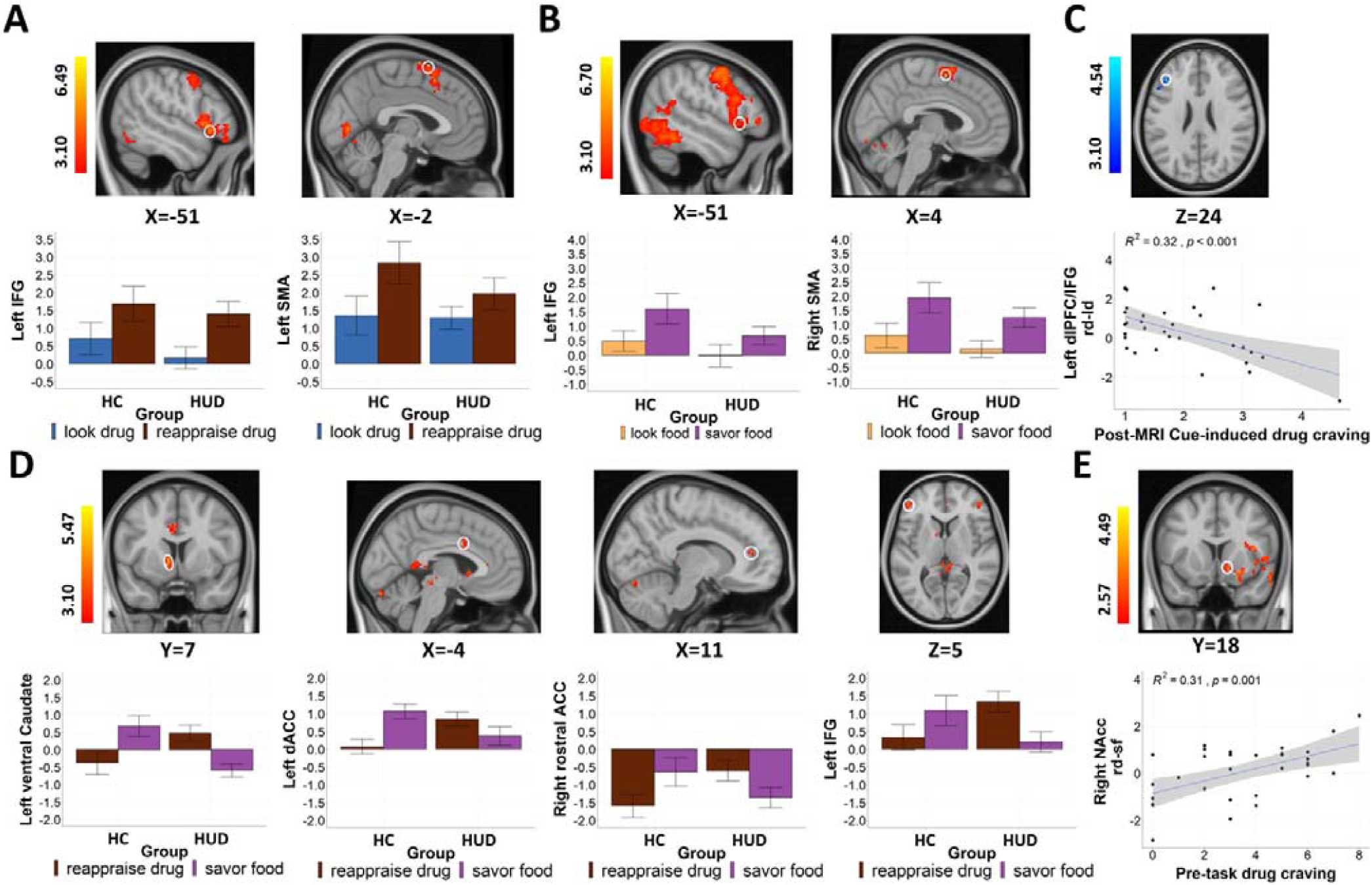
Brain activation for the reappraise and savor effects. A). Clusters indicating the effect of reappraisal across all participants, with higher activity in the left IFG and left SMA during the reappraise drug>look drug contrast. B). Clusters indicating the effect of savoring across all participants, with higher activity in the left IFG and right SMA during the savoring food>look food contrast. C) In the HUD group, significant whole-brain negative correlations between post-MRI drug cue-induced craving and activation in the left dlPFC/IFG activation during the reappraise drug>look drug contrast. D) Clusters indicating increased activity in regions including the left ventral caudate, left dACC, right rostral ACC, and left IFG in iHUD compared to HC during the reappraise drug>savor food contrast. E) Clusters indicating significant whole-brain positive correlations between pre-task baseline drug craving and activation in the right NAcc during the reappraise drug>savor food contrast in iHUD. In all panels, for visualization purposes, bar graph and scatter plots depict BOLD signal and correlation derived via 3-mm radius masks centered on coordinates from peak activity (circled in white), except in the left IFG in B to match A (see supplemental Figure 3A for peak effects). Error bars represent standard error.

## Discussion

Here we demonstrate drug cue-reactivity related hyperactivations in the NAcc, dlPFC, IFG, and vmPFC, the latter predicting post-task drug craving in iHUD as compared to HC; hypoactivations of the dlPFC and IFG while viewing non-drug rewards (food images) was also revealed in the iHUD. Although group comparisons for drug reappraisal and food savoring (vs. their respective passive viewing) did not reveal significant results, these conditions’ direct contrast revealed drug-related cortico-striatal hyperactivity (drug-reappraisal>food-savoring) in the iHUD as predicted in the striatum by pre-task baseline drug craving. Reappraisal-related increases in the dlPFC/IFG predicted lower drug cue-induced craving in the iHUD.

Increased vmPFC and NAcc reactivity to drug vs. neutral cues supports previous findings across substance use disorders (e.g., alcohol, cocaine, nicotine, and marijuana) as documented in a meta-analysis^60^, and also within iHUD specifically,^61–63^ and as compared to HC subjects.^25,26^ Recruitment of the vmPFC and NAcc may subserve processing and appraisal of stimulus salience.^64,65^ In support of this potential mechanism, we found that the higher the vmPFC drug cue-reactivity, the higher the post-task self-reported heroin craving, also in agreement with previous results (linking the vmPFC and NAcc/subcallosal gyrus with drug cravings).^18,24–26^ In a previous study, higher initial drug cue-induced NAcc/subcallosal gyrus activity characterized iHUD who relapsed within the following 3 months as compared to those who stayed abstinent,^25^ highlighting the clinical predictive relevance of these results. Consistent with another study in iHUD^30^, here we also found hyperreactivity in the dlPFC/IFG when directly comparing drug cue viewing to another salient reinforcer (food images). Such heightened drug cue reactivity as directly compared to attenuated reactivity during non-drug reward extends the iRISA model^3,4^ to iHUD.

During reappraisal of drug cues, all participants had increased activity in the IFG, dlPFC and SMA, and these increases in the dlPFC/IFG predicted lower drug cue-induced craving in iHUD. These findings are in agreement with these regions’ roles in downregulating emotional reactivity via reappraisal in the general population^37–39^ and in individuals with drug addiction.^40^ The specific underlying neural mechanism of reappraisal of drug cues has been attributed to enhanced PFC-mediated cognitive control,^66^ suggested to disrupt subcortically-driven attention bias to salient stimuli.^67^ Consistent with our results, reappraisal of cigarette cues in current cigarette smokers increased dlPFC and IFG activations with parallel decreases in cigarette craving.^33^ Based on recent studies demonstrating deficits in LPP activity during reappraisal of unpleasant cues in opioid misuse and HUD^41,68^, we expected group differences in drug cue reappraisal. Several explanations could contribute to this null effect, including: 1) different reappraisal targets used across the studies (heroin images in the current study vs. generally unpleasant/negative cues in previous studies of substance use disorders^68,69^); and 2) the current iHUD were all receiving medication assisted treatment and highly motivated to maintain abstinence. Their ability to recruit cognitive control regions during reappraisal at a similar level to that of HC is potentially indicative of an intact ability to mobilize higher-order executive functions while adopting specific reappraisal instructions for the duration of this task. The generalization of this ability during spontaneous, non-instructed potent drug cue encounters, or under prolonged challenges and/or stress, and as relevant to the regulation of real-life craving, remains to be ascertained.

In addition to the drug cue reactivity and its top-down regulation during fMRI, for the first time in heroin addiction we included non-drug reward (food) savoring. Similar to the absence of group differences during drug reappraisal, the groups did not differ during this condition, suggesting comparable abilities in iHUD to recruit the effort (cortically- and subcortically-mediated) needed for this alternative reward processing. Importantly, when directly contrasting these two conditions, iHUD showed increased activations during drug cue reappraisal while HC showed similar increases during food cue savoring across our cortico-striatal regions of interest encompassing the ventral caudate, the ACC and IFG. The enhanced activation in the ventral caudate is consistent with the shift from hedonic (ventral striatal) to habit (dorsal striatal) responses,^70^ although the caudate also has a role in cognitive reappraisal^71^ via its functions in goal-directed control, including selecting appropriate sub-goals and initiating/executing correct action schemas.^72^ In our study, baseline drug craving (measured before the MRI task) correlated with NAcc activity during reappraisal vs. savoring in iHUD, highlighting a potential state that facilitates such an imbalance (drug reappraisal>food savoring). Taken together, these results suggest that among treatment-seeking iHUD, the resources needed for reappraisal of drug cues may outweigh those recruited during savoring of non-drug rewards in regions comprising the reward, habit, salience and cognitive control networks. Re-balancing of reappraisal and savoring may be required for adaptive hedonic regulation and recovery in addiction.

Several study limitations should be considered. First, all iHUD were recruited from a medication assisted treatment program. Considering that methadone-maintained iHUD have displayed enhanced drug-cue related mesocorticolimbic activity compared to those in protracted abstinence,^75^ similar investigations should be extended to both abstinent and non-treatment seeking drug using iHUD. Longitudinal investigations are needed to inspect the cortico-striatal signature of drug cue-reactivity and its regulation over the course of the drug addiction cycle and recovery in iHUD. The iHUD and HC samples significantly differed in education, verbal IQ, and self-reported depression. Although these variables were not associated with our outcomes of interest and thus could not have substantially affected our results, larger and more closely matched samples can better inform the contributions to results of individual differences in these and other demographic variables. Lastly, the samples also differed on cigarette smoking. Although there were no correlations between FTND/number of cigarettes smoked per day and our outcome variables within iHUD, suggesting cigarettes smoking did not contribute to our results, future studies should recruit HC participants who are cigarette smokers.

To the best of our knowledge, this is the first neuroimaging study to compare iHUD and HC on cortico-striatal markers of cue-reactivity and its modulation via reappraisal and savoring. Congruent with the iRISA model, cortico-striatal hyperreactivity to drug (at the expense of food) cues, and the imbalanced reappraisal and savoring responses in iHUD (which correlated with drug craving), suggest an attentional bias to drug cues at the expense of processing alternative rewards. These results provide insight for cognitive-behavioral treatments including reappraisal and savoring training (e.g., as those integrated in MORE^47^), as well as non-invasive neuromodulation targeting reward, salience, and cognitive control networks to reduce craving and enhance recovery in iHUD.

## Data Availability

All data produced in the present study are available upon reasonable request to the authors

