## Supplementary materials for "Cortico-striatal engagement during cue-reactivity, reappraisal, and savoring of drug and non-drug stimuli predicts craving in heroin addiction"

**eAppendix**

**Exclusion criteria**

Exclusion criteria for all participants were the following: 1) DSM-5 diagnosis for schizophrenia or developmental disorder; 2) Head trauma with loss of consciousness (>30 min); 3) History of neurological disease of central origin; 4) Cardiovascular, metabolic, endocrinological, oncological, autoimmune, and infectious diseases common in iHUD including Hepatitis B and C or HIV/AIDS; 5) Metal implants or other MR contraindications (including pregnancy). We did not exclude for DSM-5 diagnosis of a drug use disorder other than opiates as long as heroin was the primary drug of choice/reason for treatment-seeking since iHUD commonly use alcohol, amphetamines, benzodiazepines, other sedatives, cocaine, and marijuana. Exclusion criteria for the HC were the same, except history of any drug use disorder was prohibitive.

**Comorbidities**

Other comorbidities in the iHUD included cocaine use disorder (n=8), sedative use disorder (n=4), general anxiety disorder (n=4), alcohol use disorder (n=3), persistent depressive disorder (n=2), major depressive disorder (n=2), meth/amphetamine use disorder (n=2), panic disorder (n=1), marijuana use disorder (n = 1), antisocial personality disorder (n=1), and specific phobia (n=1). These comorbidities were either in partial or sustained remission at time of study. No comorbidities were found for the HC subjects.

**Task-related instructions**

During the look condition, participants were instructed to “keep viewing the picture normally”. During the reappraise condition, participants were instructed to reduce their emotional reactivity to the heroin pictures in three practice trials, each providing a different strategy: 1) “Try to imagine that the scenario is not real, that it is from a movie, and these are all actors”; 2) “Try to imagine that the heroin is not real, that it is just a prop”; 3) “You can focus on how this is just a picture, and tell yourself that it is not real heroin”. During the savor condition, participants were instructed to increase their emotional reactivity to the food pictures in three practice trials, each providing a different savoring strategy: 1) “You can imagine that you are holding the food in the picture, and feeling the weight of it in your hands, enjoying its pleasant smell”; 2) “You can focus on how good the food looks or imagine how good it would taste, savoring the delicious taste of the food”; 3) “Imagine the sensation of how it would feel in your mouth or how it feels once you’ve eaten it”. Participants were instructed to verbally describe their reappraisal and savoring strategies during these practice trials to ensure task comprehension, but they were instructed to refrain from speaking during the fMRI task trials in the scanner.

**Pre- and post-cue reactivity task ratings**

Participants provided heroin and food craving ratings, as well as a rating of motivation to complete the task immediately before the fMRI cue-reactivity task (i.e., “Please rate how strong your desire for heroin is currently on a scale of 0-9”, “Please rate how strong your desire for food is currently on a scale of 0-9”, and “Please rate your motivation to complete this task on a scale of 0-9”). Immediately after the cue-reactivity task, in addition to the same three questions, participants were asked to provide self-evaluation of the difficulty and effectiveness of their reappraisal and savoring performance on a 10-point scale (i.e., “How difficult did you find it to decrease your emotional reactivity to the heroin pictures during this task?”, “How well do you think you decreased your emotional reactivity to the heroin pictures during this task?”, “How difficult did you find it to increase your emotional reactivity to the food pictures during this task?” and “How well do you think you increased your emotional reactivity to the food pictures during this task?”). Means and standard errors of each rating for both groups are plotted in eFigure 1.

To analyze pre- and post-task drug/food craving ratings between groups, a 2 (group: iHUD/HC) by 2 (time: pre/post) by 2 (image: drug/food) mixed ANOVA was conducted. We found significant main effects of group [iHUD>HC: F(1, 51) = 49.81, p < 0.01] and image [food>drug: F(1, 51) = 91.16, p < 0.01] and a significant group × image interaction [F(1, 51) = 7.82, p < 0.01]. Follow up analyses showed that the iHUD group had higher drug craving ratings than the HC group [t(51) = 7.99, p < 0.01]. The time main effect [F(1, 51) = 3.42, p = 0.07] and the group × time interaction [F(1, 51) = 0.09, p = 0.76], time × image interaction [F(1, 51) = 0.97, p = 0.33], and group × time × image interaction [F(1, 51) = 0.02, p = 0.89] were not significant. A 2 (group: iHUD/HC) by 2 (time: pre/post) mixed ANOVA for motivation ratings showed neither significant main effects of group [F(1, 51) = 0.27, p = 0.61], time [F(1, 51) = 0.09, p = 0.77], nor an interaction between group and time [F(1, 51) = 0.04, p = 0.85]. Regarding self-evaluated reappraisal and savoring performance, two separate 2 (group: iHUD/HC) by 2 (image: drug/food) mixed ANOVAs for difficulty and effectiveness were conducted. There were no significant group main effects [difficulty: F(1, 51) = 1.85, p = 0.18, effectiveness: F(1, 51) = 1.39, p = 0.24], image main effects [difficulty: F(1, 51) = 0.40, p = 0.53, effectiveness: F(1, 51) = 0.54, p = 0.47] nor group × image interaction effects [difficulty: F(1, 51) = 0.27, p = 0.61, effectiveness: F(1, 51) = 1.32, p = 0.26]. Overall, these results suggest that the HUD group had higher heroin craving than the HC group, but that there were no group differences in food craving, motivation, or self-evaluated reappraisal/savoring performance nor task induced changes in any of these ratings.

**Post-MRI picture ratings**

After the MRI, participants provided image-specific valence and arousal ratings on a subset of the drug, food, and neutral images, as well as wanting ratings on the drug and food images presented in the task. The images to be rated were pseudorandomized across participants. For valence ratings, participants were asked ‘How pleasant do you find the above picture?’ on a scale from 1 (very unpleasant) to 5 (very pleasant). For arousal, participants were asked “How emotional do you feel about the above picture?” on a scale from 1 (calm, no emotion) to 5 (extremely emotional). For wanting, participants were asked “How strong is your desire to use the above substance (or food)?” on a scale from 1 (no desire) to 5 (extreme desire). Means and standard errors of each rating for both groups are plotted in eFigure 2.

To test potential differences between valence and arousal ratings for food, drug and neutral images, two separate 2 (group: iHUD/HC) by 3 (images: drug/food/neutral) mixed ANOVAs were conducted for valence and arousal. For valence, there was a significant main effect of image [food>neutral>drug: F(1.30, 66.43) = 163.51, p < 0.01]. The main effect of group [F(1, 51) = 0.02, p = 0.88] and the group × image interaction [F(1.30, 66.43) = 2.21, p = 0.14] did not reach significance. For arousal, the main effects of group [iHUD>HC: F(1, 51) = 7.85, p < 0.01] and image [food>drug>neutral: F(1.51, 76.93) = 9.8, p < 0.01] were significant while their interaction was not [F((1.51, 76.93) = 0.26, p = 0.71]. A 2 (group: iHUD/HC) by 2 (image: drug/food) mixed ANOVA on the wanting ratings showed significant main effects of group [iHUD>HC: F(1, 51) = 11.47, p < 0.01] and image [food>drug: F(1, 51) = 62.08, p < 0.01] and a significant group × image interaction [F(1, 51) = 4.13, p = 0.047] explained by significantly higher drug wanting ratings in the HUD than the HC group [t(51) = 4.34, p < 0.01], consistent with the results for craving reported above.

**MRI data acquisition and preprocessing**

The MRI protocol was optimized to be Human Connectome Project compatible^1^. The blood-oxygen-level-dependent (BOLD) fMRI responses were measured as a function of time using T2*-weighted single-shot multiband accelerated gradient-echo echo-planar image (EPI) sequence [TE/TR=35/1000 ms, 2.1 isotropic mm resolution, 70 axial slices with no gap for the whole brain (147mm) coverage, FOV 206 × 181 mm, matrix size 96 × 84, 60°-flip angle (approximately Ernst angle), multi-band factor of 7, blipped CAIPIRINHA phase-encoding shift=FOV/3, 1860 kHz/Pixel bandwidth with ramp sampling, echo spacing 0.68 ms, and echo train length 57.1 ms]. T1-weighted structural images were acquired using a 3D MPRAGE sequence [FOV 256 × 256 × 180 mm^3^, 0.8 mm isotropic resolution, TR/TE/TI = 2400/2.07/1000 ms, flip angle 8° with binomial (1, −1) fat saturation, bandwidth 240 Hz/pixel, echo spacing 7.6 ms, and in-plane acceleration (GRAPPA) factor of 2, with a total acquisition time of 7m 2s.

Raw data were converted via HeuDiConv (<https://github.com/nipy/heudiconv>) and adapted to the Brain Imaging Data Structure standards^2^ for portability and reproducibility and preprocessed using the Nipype-based fMRIprep pipeline (version 20.2.1).^3,4^ The structural images were corrected for intensity non-uniformity and skull stripped with ANTs^5,6^. Brain tissue was segmented into the cerebrospinal fluid, white matter, and gray matter through FSL’s FAST^7^. Volume-based spatial normalization through nonlinear registration into ICBM 152 nonlinear asymmetrical template was performed with ANTs^5,8^. The functional images were corrected for distortion using echo-planar field maps acquired in opposing phase-encoding directions via 3dQwrap in AFNI^9^. Motion artifacts were estimated and corrected via FSL’s MCFLIRT^10^. Motion- and distortion- corrected images were then co-registered to participants’ T1w images with the boundary-based registration with 9 degrees of freedom using FSL’S FLIRT^8,11^, and normalized to ICBM 152 nonlinear asymmetrical template^8^. Data were visually inspected and those deemed poorly-registered underwent Freesurfer’s *recon-*all^12^ prior to repeating fMRIprep’s registration pipeline. In addition to fMRIprep, we identified volumes with spikes in translation and rotation parameters using a typical boxplot threshold (75th percentile + 1.5 times the interquartile range) in relation to a reference image volume using FSL’s *fsl_motion_outlier*. The identified outlier time points were used as regressors of no interest at the run-level general linear model (GLM). On average, we regressed out 5.30% of the total volumes in each run (range: 0.5%-14.89%). The HUD (mean = 23.59±10.77) group displayed significantly more outlier time points than the HC (mean = 17.89±8.41) group [t(51) = 2.05, p = 0.046]; however, no significant whole-brain correlation was found between the number of outlier time points and the peak signal in the contrasts of interest across all participants. A high-pass filter (100 s cutoff) was applied to the functional data to ignore scanner drift. Lastly, the preprocessed data were spatially smoothed with a Gaussian kernel (5 mm full-width at half maximum) to improve signal to noise ratio.

Parameter estimates were generated for each participant using the GLM approach via FSL’s FEAT (version 6.0.0).^13^ To control for the potential effects of passive viewing, we created an additional contrast (reappraise-drug minus look-drug>savor-food minus look-food). Ratings for the whole-brain correlations analyses were entered as regressors in separate models to reveal potential relationships between self-reported general and cue-induced drug craving and neural signaling associated with cue-reactivity (look drug>look neutral; look drug>look food), reappraisal (reappraise drug>look drug; reappraise drug>savor food), and savoring (savor food>look food).

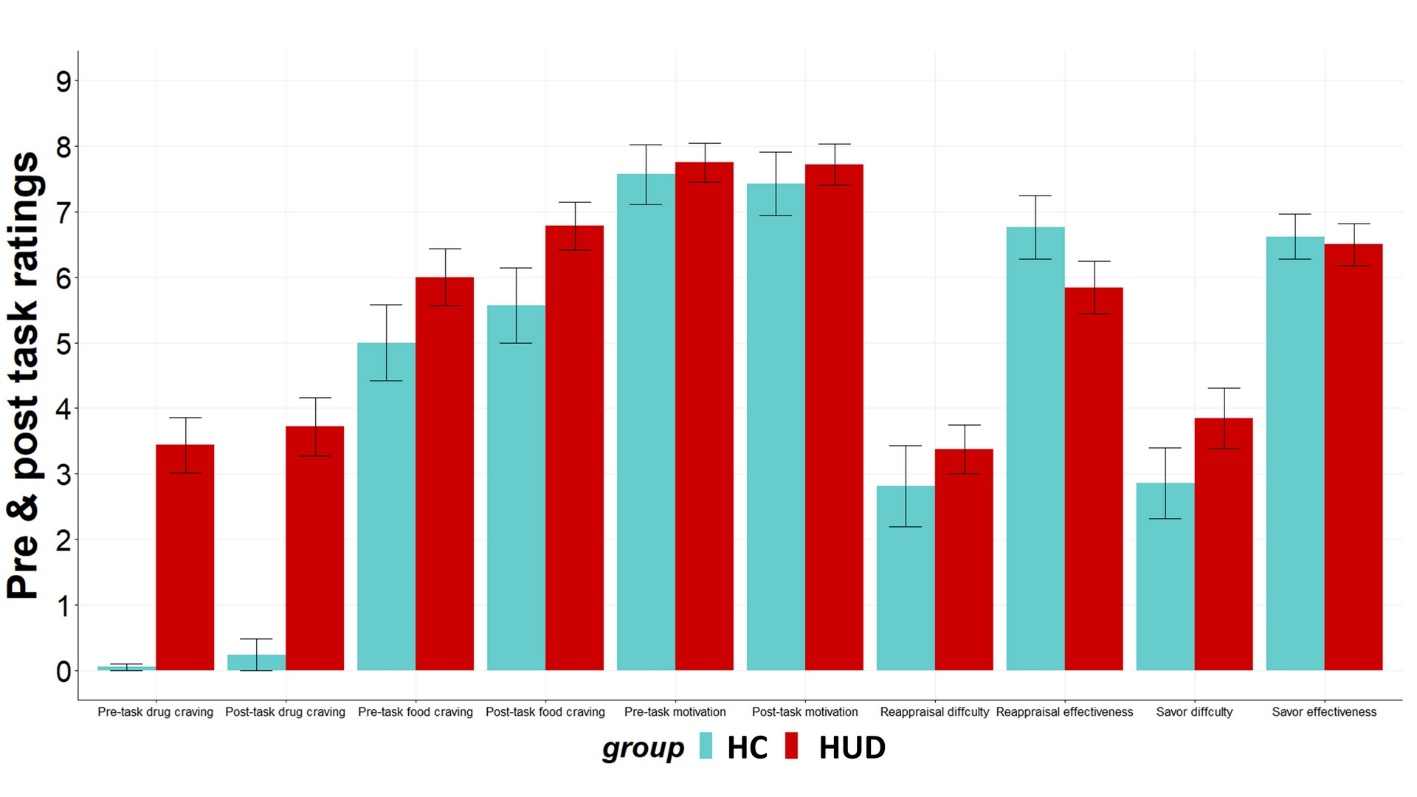

**eFigure 1.** Bar plots for pre- and post-task ratings. Error bars represent standard error

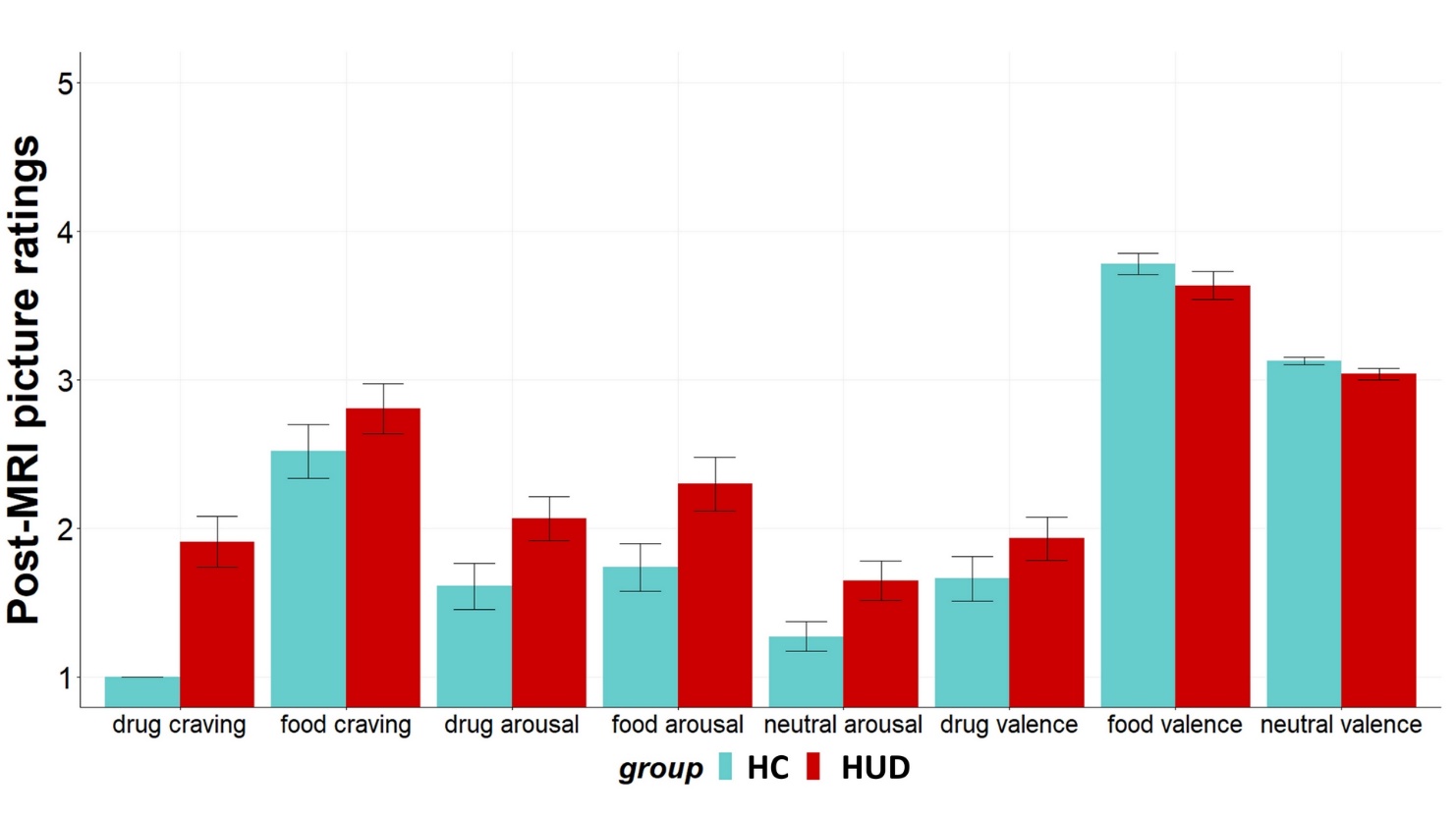

**eFigure 2.** Bar plots for post-MRI picture ratings. Error bars represent standard error

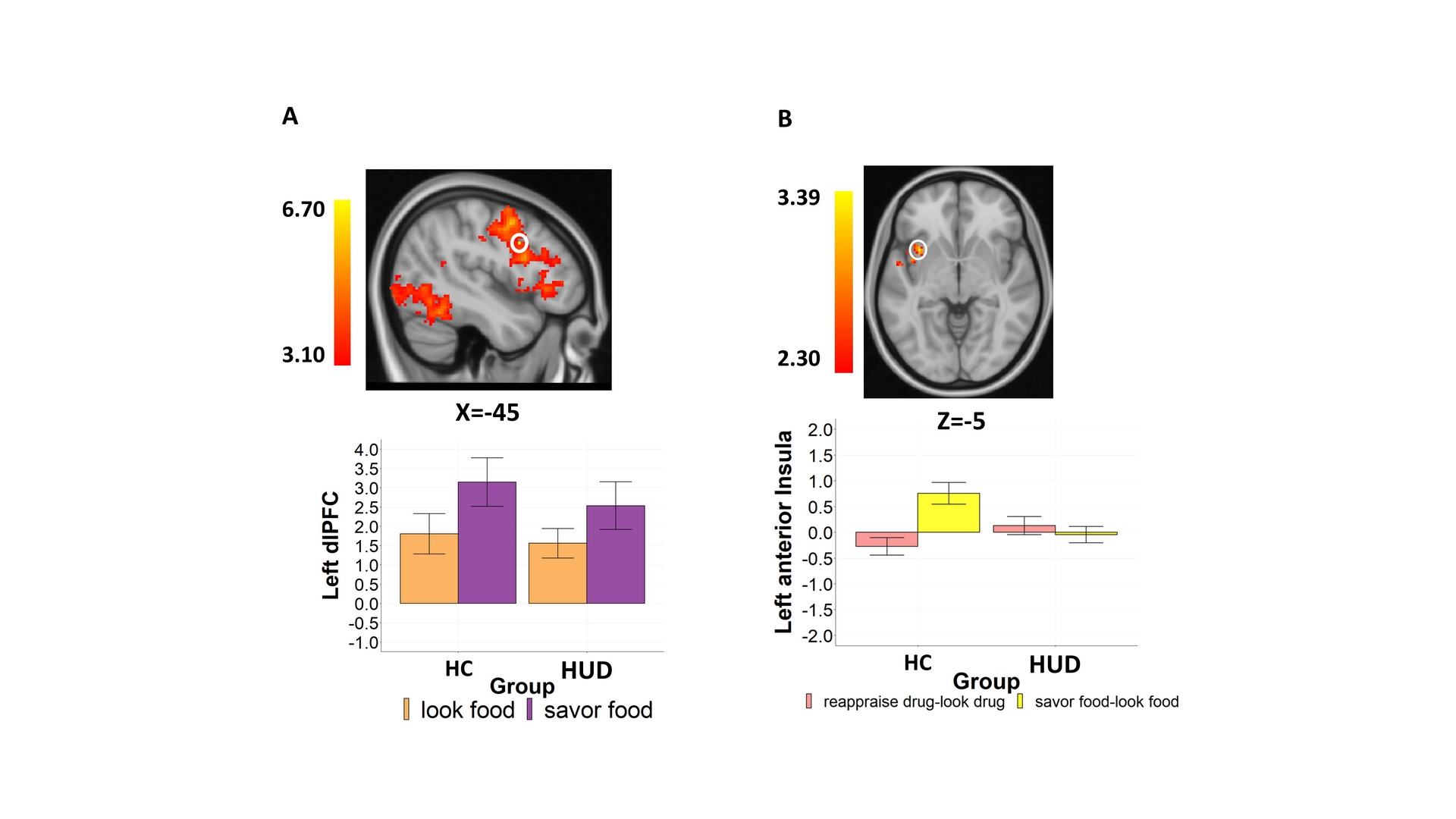

**eFigure 3.** Between group comparison for savor food>look food and reappraise drug-look drug>savor food-look food. For visualization purposes, bar graph and scatterplots depict BOLD signal and correlation derived via 3-mm radius masks centered on coordinates from peak activity (circled in white). Error bars represent standard error.

**eTable 1. Clusters for between-groups direct contrasts**

|  | **Side** | **Voxels** | **Peak Z** | **X** | **Y** | **Z** | **BA** |
| --- | --- | --- | --- | --- | --- | --- | --- |
| **Drug cue-reactivity**  iHUD>HC: reappraise drug minus look drug>savor food minus look food (Z>2.3) | | | | | | | |
| Anterior insula | L | 301 | 3.35 | -34 | 11 | -5 | - |

**eTable 2. Whole-brain drug craving correlations in iHUD**

|  | **Side** | **Voxels** | **Peak Z** | **X** | **Y** | **Z** | **BA** |
| --- | --- | --- | --- | --- | --- | --- | --- |
| look drug>neutral with post-MRI drug cue-induced craving | | | | | | | |
| Posterior cingulate cortex | L&R | 55 | +4.03 | 0 | -49 | 31 | BA23 |
| look drug>food with post-MRI drug cue-induced craving | | | | | | | |
| Posterior cingulate cortex | L | 79 | +4.1 | -2 | -45 | 29 | BA23 |
| Precuneus | R | 76 | +4.56 | 4 | -60 | 73 | BA7 |
| Superior parietal lobule | L | 446 | +5.11 | -19 | -64 | 54 | BA7 |
| Inferior temporal gyrus | R | 92 | -4.11 | 56 | -21 | -26 | BA20 |
| look drug>food with post-task drug craving | | | | | | | |
| Middle occipital gyrus | R | 66 | +4.04 | 54 | -66 | 27 | BA39 |
| reappraise drug>look drug with pre-task drug craving | | | | | | | |
| Inferior parietal lobule | L | 67 | -3.89 | -49 | -23 | 37 | BA40 |
| reappraise drug>savor food with post-task drug craving | | | | | | | |
| Intracalcarine cortex | R | 64 | +4.09 | 11 | -60 | 16 | BA18 |
| reappraise drug>savor food with post-MRI drug cue-induced craving | | | | | | | |
| Precuneus | R | 52 | +4.15 | 11 | -77 | 60 | BA7 |
| Superior occipital gyrus | R | 76 | +3.93 | 22 | -77 | 20 | BA19 |
| Insula | L | 54 | -4.04 | -36 | -10 | 14 | - |

A positive peak Z designates a positive correlation while a negative peak Z designates a negative correlation
